# Clinical Epidemiological Features and Risk Factor Weight Remodeling in Gallstone Patients on the Plateau

**DOI:** 10.64898/2026.08.16.26360533

**Authors:** Zhiqiang Wang, Guoliang Ren, Zhongfeng Dang, Wei Su, Yabing Ma, Ping Li, Dongde Ji, Liansheng Li, Junlin Gao

**Affiliations:** Department of Hepatobiliary and Pancreatic Surgery, Qinghai Red Cross Hospital, Xining, Qinghai 810000, China; Ultrasound and Electrocardiography Center, Gansu Hospital of Sun Yat-sen University Cancer Center, Lanzhou, Gansu 730050, China

**Keywords:** Gallstones, Cholecystitis, Plateau, Epidemiology, Systolic blood pressure, Risk factors

## Abstract

**Objective:** To test the hypothesis that body mass index (BMI) replaces sex as the core risk factor for cholecystitis in plateau populations, and to characterize the true risk factor profile of gallstone patients at high altitude.

**Methods:** A single-center retrospective cohort study included 605 elective laparoscopic cholecystectomy patients at Qinghai Red Cross Hospital (2,260 m; 2020-2023), categorized into simple gallstones (n=434) and gallstones with cholecystitis (n=171). Univariate analysis, multivariate logistic regression, nested model comparison, interaction analysis, and sensitivity analyses were performed. All statistics were independently recomputed using Python 3.11 and cross-validated against original statistical deliverables.

**Results:** The original hypothesis was falsified. The cohort had a mean age of 43.7±11.8 years, BMI of 24.2±3.8 kg/m², female-to-male ratio of 2.10:1, and SBP of 117.4±15.8 mmHg. Multivariate logistic regression (adjusting for age, BMI categories, sex, SBP, and DBP) showed that SBP was the only significant positive predictor (OR=1.027/mmHg, 95%CI: 1.008-1.047, P=0.005), while BMI overweight (OR=1.038, P=0.856) and obesity (OR=0.645, P=0.137) were non-significant, as was sex (OR=0.814, P=0.322). Age showed a significant negative association (OR=0.978/year, P=0.007), constituting an “age paradox” with younger patients having higher cholecystitis rates (<30 years: 36.8% vs ≥60 years: 25.0%; Spearman ρ=-0.087, P=0.032), which may reflect selection bias or plateau-specific mechanisms. Nested model comparison showed that adding blood pressure to the classical model (age+BMI+sex, AUC=0.575) significantly improved discrimination (AUC=0.615, ΔAUC=+0.040, 95%CI: +0.007 to +0.084; LR χ²=8.19, P=0.017). SBP was non-significant in univariate analysis (OR=1.005, P=0.346) due to a suppression effect: age was a negative confounder, simultaneously increasing SBP and decreasing cholecystitis risk, thereby masking the true SBP effect.

**Conclusion:** The risk weight of cholecystitis is remodeled in the plateau hypoxic environment, but in a direction opposite to the original hypothesis: SBP is the only significant positive predictor (P=0.049 in the SBP-only recommended model), while BMI and sex are non-significant, and age shows an inverse association. Blood pressure management should be integrated into the risk stratification system for plateau cholecystitis. The original hypothesis that BMI replaces sex is explicitly falsified.

## Introduction

Gallstones are a highly prevalent benign biliary disease worldwide, and their occurrence and progression are jointly regulated by multiple factors including age, sex, body weight, and geographical environment [1,2]. Plain areas have established the classic epidemiological paradigm of female predominance and gender-dominant pathogenesis [3]. However, the unique hypoxic environment and distinctive metabolic characteristics of high-altitude regions can significantly reshape physiological homeostasis [4], potentially altering the traditional pathogenic weight distribution of biliary diseases.

In preliminary theoretical reasoning, our research team proposed the working hypothesis that “BMI replaces sex as the core risk factor for cholecystitis at high altitude,” based on the rationale that the hypoxic plateau environment induces insulin resistance and lipid metabolism disorders [4], amplifying the pro-inflammatory effect of BMI on gallbladder inflammation and thereby replacing sex as the core risk factor. However, this hypothesis has not been rigorously tested with large-sample raw data. The credibility of a scientific hypothesis depends on its falsifiability and its survival when confronted with raw data; when systematic deviations arise between predicted and observed data, the hypothesis should be revised based on real data rather than being preserved [5].

This study systematically tests the above working hypothesis based on raw clinical data from 605 plateau gallstone patients, and re-characterizes the true risk factor profile of plateau cholecystitis under a data-driven revision framework. This study is based on the same clinical cohort as Series I (surgical efficiency analysis) and Series III (health economics analysis), but focuses on an independent scientific question: Series I examines surgical efficiency heterogeneity, this study investigates disease risk-factor remodeling, and Series III develops cost control models. There is no risk of duplicate publication among the three.

### Research contributions

This study makes the following three specific contributions to the literature:

First, this study supplements epidemiological baseline data for gallstone patients in the Qinghai-Tibet Plateau region (n=605), enriching the epidemiological evidence for plateau biliary diseases.

Second, through full recomputation of raw data, it explicitly falsifies the original “BMI replaces sex” working hypothesis, and revises the risk factor weight direction to “vascular factors supersede metabolic/demographic factors” based on real data.

Third, it reveals the age paradox and systolic blood pressure suppression effect in plateau cholecystitis, providing new evidence-based support for risk stratification of plateau biliary diseases.

## Methods

### Study design and setting

This single-center retrospective cohort study was conducted at Qinghai Red Cross Hospital (Xining, Qinghai, China; altitude 2,260 m). The study was approved by the Ethics Committee of the hospital (Approval No.: LW-2026-71) and followed the principles of the Declaration of Helsinki [6]. The requirement for informed consent was waived due to the retrospective and anonymous nature of the data. This study used the same inclusion, exclusion, and ethical standards as Series I. From January 2020 to December 2023, 613 cases were initially enrolled. After excluding invalid records, 605 elective laparoscopic cholecystectomy (LC) patients were ultimately included. The sample size differences across studies (Series I: n=591; this study and Series III: n=605) reflect different analytical objectives: Series I focused on surgeon efficiency comparison and only included cases from the top 7 surgeons by volume; this study and Series III focused on patient-level analysis and included all eligible cases.

### Data collection

Complete clinical data were systematically extracted from the hospital’s electronic medical record system. The main variables included: (1) Demographic baseline data: age, sex, height, weight, and body mass index (BMI); (2) Clinical baseline indicators: systolic blood pressure (SBP), diastolic blood pressure (DBP), pulse pressure (PP=SBP-DBP), and pre-operative diagnosis classification (simple gallstones vs. gallstones with cholecystitis); (3) Comorbidities: hypertension. All study data were independently entered by two researchers and cross-checked. All 605 patients included had complete analysis variables with no missing data.

### Definitions and measurements

BMI was categorized according to the standards recommended by the Working Group on Obesity in China [7]: underweight (<18.5 kg/m²), normal (18.5-23.9 kg/m²), overweight (24.0-27.9 kg/m²), and obese (≥28 kg/m²). Hypertension was defined as systolic blood pressure ≥140 mmHg and/or diastolic blood pressure ≥90 mmHg. The study population was divided into the simple gallstones group and the gallstones with acute/chronic cholecystitis group to compare baseline differences and identify risk factors for cholecystitis progression.

### Statistical analysis

All statistical analyses were performed using Python 3.11 (pandas 2.0, statsmodels 0.14, scipy 1.11, scikit-learn 1.3), with cross-validation against SPSS 26.0. Continuous variables are presented as mean±standard deviation and compared using independent samples t-tests; categorical variables are expressed as n (%) and compared using chi-square tests. Binary logistic regression was performed with “cholecystitis” as the binary dependent variable, including age, BMI categories (overweight, obese, with normal as reference), sex, SBP, and DBP. Odds ratios (OR), 95% confidence intervals (CI), and P values were calculated, with significance level α=0.05.

Due to high collinearity between SBP and DBP (VIF>100), the main model adopted an SBP-only approach to avoid collinearity interference, with the full model (SBP+DBP) reported as a comparator. The Hosmer-Lemeshow test was used to assess goodness-of-fit, and the C-statistic (AUC) was used to assess discrimination.

Nested model comparison: Model A was the classical model (age+BMI categories+sex), and Model B was the classical model+SBP+DBP. The likelihood ratio test (LR test) was used to compare model fit, and Bootstrap (n=1000) was used to calculate the 95% CI of ΔAUC.

Interaction analysis: SBP×sex, SBP×age group (<40 vs ≥40), SBP×BMI, and BMI×sex interaction terms were constructed to test for effect modification.

Sensitivity analysis: (1) Rebuild the model after excluding the underweight group (n=23); (2) Model separately by age strata (<40 vs ≥40); (3) Model separately by sex.

Sample size calculation was performed a priori using G*Power 3.1.9.7 (F tests → Logistic regression). With the effect of SBP on cholecystitis progression as the primary hypothesis, parameters were set as: expected OR=1.3, α=0.05 (two-tailed), 1-β=0.80, number of predictors k=5. The required minimum number of events was 50, with a minimum total sample size of 200. The actual study included 605 patients with 171 cholecystitis events, yielding an events-per-variable (EPV) ratio of 171/5=34.2, satisfying the logistic regression requirement (EPV≥10) [8] and providing adequate statistical power.

This study followed the STROBE (Strengthening the Reporting of Observational Studies in Epidemiology) reporting guidelines [9]; the checklist is provided in the supplementary materials.

## Results

### Baseline characteristics of the entire cohort

A total of 605 patients were included in this study, including 195 males (32.2%) and 410 females (67.8%), yielding a female-to-male ratio of 2.10:1. The mean age was 43.7±11.8 years. Age stratification: <40 years 236 cases (39.0%), 40-60 years 329 cases (54.4%), >60 years 40 cases (6.6%), indicating a predominantly middle-aged study population.

The mean BMI of the entire cohort was 24.2±3.8 kg/m², SBP 117.4±15.8 mmHg, DBP 77.4±11.0 mmHg; hypertension was present in 102 patients (16.9%). BMI stratification: underweight 23 (3.8%), normal 274 (45.3%), overweight 220 (36.4%), obese 88 (14.5%), with 50.9% of patients having overweight or obesity.

Diagnosis groups: simple gallstones 434 (71.7%), gallstones with cholecystitis 171 (28.3%).

**Table 1.** Baseline characteristics of the entire cohort (N=605)

| Characteristic | Value |
| --- | --- |
| Age (years), mean±SD | 43.7±11.8 |
| Sex, n (%) |  |
| Male | 195 (32.2) |
| Female | 410 (67.8) |
| Female:Male ratio | 2.10:1 |
| Age strata (years), n (%) |  |
| <40 | 236 (39.0) |
| 40-60 | 329 (54.4) |
| >60 | 40 (6.6) |
| BMI (kg/m<sup>2</sup>), mean±SD | 24.2±3.8 |
| BMI categories, n (%) |  |
| Underweight (<18.5) | 23 (3.8) |
| Normal (18.5-23.9) | 274 (45.3) |
| Overweight (24.0-27.9) | 220 (36.4) |
| Obese ($\geq 28.0$) | 88 (14.5) |
| Overweight+Obese ($\geq 24.0$) | 308 (50.9) |
| SBP (mmHg), mean±SD | 117.4±15.8 |
| DBP (mmHg), mean±SD | 77.4±11.0 |
| Hypertension, n (%) | 102 (16.9) |
| Diagnosis group, n (%) |  |
| Simple gallstones | 434 (71.7) |
| Gallstones with cholecystitis | 171 (28.3) |

### Univariate comparison between groups

Among the 605 patients, 434 were in the simple gallstones group and 171 in the gallstones with cholecystitis group. Univariate analysis showed that only age differed significantly between groups (P=0.029), and the direction was opposite to the classic understanding: the cholecystitis group was younger (42.0±12.3 vs 44.3±11.6 years). BMI, SBP, DBP, sex, and hypertension showed no significant differences between groups (P>0.05). See Table 2.

**Table 2.** Baseline characteristics comparison between the simple gallstones group and the gallstones with cholecystitis group.

| Characteristic | Simple gallstones<br>(n=434) | Gallstones with<br>cholecystitis<br>(n=171) | Statistical test | P value |
| --- | --- | --- | --- | --- |
| Age (years) | 44.3±11.6 | 42.0±12.3 | t=2.19 | 0.029 |
| BMI (kg/m<sup>2</sup>) | 24.4±3.9 | 23.9±3.5 | t=1.48 | 0.139 |
| Gender (M/F) | 146/288<br>(33.6%/66.4%) | 49/122<br>(28.7%/71.3%) | $\chi^2=1.18$ | 0.278 |
| SBP (mmHg) | 117.0±15.6 | 118.3±16.1 | t=-0.94 | 0.346 |
| DBP (mmHg) | 77.6±10.4 | 76.8±12.5 | t=0.79 | 0.430 |
| Hypertension<br>[n(%)] | 74 (17.1) | 28 (16.4) | $\chi^2=0.01$ | 0.937 |

Note: Continuous variables were compared using independent samples t-tests; categorical variables were compared using chi-square tests. Age differed significantly between groups (P=0.029), and the direction was opposite to the classic understanding: the cholecystitis group was younger.

This univariate result already poses a preliminary challenge to the original hypothesis: if BMI were indeed the core risk factor, between-group differences should be apparent at the univariate level, but this was not observed.

### Multivariate independent risk factor analysis for cholecystitis

After binary logistic regression adjustment, the full model (SBP+DBP) showed:

SBP was the only significant positive predictor (OR=1.027/mmHg, 95%CI: 1.008-1.047, P=0.005), meaning that for each 1 mmHg increase in systolic blood pressure, cholecystitis risk increased by 2.7%.

Age showed a significant negative association (OR=0.978/year, 95%CI: 0.963-0.994, P=0.007), meaning that for each 1-year increase in age, cholecystitis risk decreased by 2.2%.

BMI overweight (OR=1.038, 95%CI: 0.695-1.551, P=0.856) and obese (OR=0.645, 95%CI: 0.362-1.149, P=0.137) were both non-significant, as was sex (OR=0.814, 95%CI: 0.541-1.224, P=0.322). DBP showed a marginally significant negative association (OR=0.974, 95%CI: 0.949-0.999, P=0.042), but given its high collinearity with SBP (VIF_SBP=107.5, VIF_DBP=100.1), its independent effect cannot be reliably estimated.

The Hosmer-Lemeshow test P=0.090 indicated good fit; C-statistic=0.615; Pseudo R²=0.023.

Given the severe collinearity between SBP and DBP (full model VIF_SBP=107.5, VIF_DBP=100.1), the recommended main model adopted an SBP-only approach (VIF_SBP=16.7, age VIF=15.4, both acceptable). Recommended main model results: SBP OR=1.013/mmHg (95%CI: 1.000-1.025, P=0.049), age OR=0.979/year (95%CI: 0.963-0.994, P=0.008), BMI overweight OR=1.011 (P=0.959), BMI obese OR=0.625 (P=0.110), sex OR=0.772 (P=0.209); AUC=0.591, H-L test P=0.822. See Table 3.

**Table 3.** Multivariate logistic regression analysis of cholecystitis.

| Variable | OR | 95% CI | P value |
| --- | --- | --- | --- |
| Full model (SBP+DBP) |  |  |  |
| Age (per 1-year increase) | 0.978 | 0.963-0.994 | 0.007 |
| BMI categories |  |  |  |
| Normal (reference) | 1.00 | — | — |
| Overweight | 1.038 | 0.695-1.551 | 0.856 |
| Obese | 0.645 | 0.362-1.149 | 0.137 |
| Sex (male=1) | 0.814 | 0.541-1.224 | 0.322 |
| SBP (per 1 mmHg) | 1.027 | 1.008-1.047 | 0.005 |
| DBP (per 1 mmHg) | 0.974 | 0.949-0.999 | 0.042 |
| Recommended main model (SBP only) |  |  |  |
| Age (per 1-year increase) | 0.979 | 0.963-0.994 | 0.008 |
| BMI categories |  |  |  |
| Normal (reference) | 1.00 | — | — |
| Overweight | 1.011 | 0.678-1.507 | 0.959 |
| Obese | 0.625 | 0.351-1.113 | 0.110 |
| Sex (male=1) | 0.772 | 0.516-1.156 | 0.209 |
| SBP (per 1 mmHg) | 1.013 | 1.000-1.025 | 0.049 |

Full model: H-L test $\chi^2=13.706$ , df=8, P=0.090; C-statistic=0.615; Pseudo R<sup>2</sup>=0.023.
Recommended main model: H-L test $\chi^2=4.371$ , P=0.822; C-statistic=0.591; Pseudo R<sup>2</sup>=0.017.
Note: In the full model, VIF for SBP and DBP were 107.5 and 100.1 respectively, indicating severe collinearity; therefore the recommended main model adopts an SBP-only approach. In the recommended main model, SBP VIF=16.7 and age VIF=15.4, slightly high but within acceptable range (<20).

Pulse pressure (PP) had a univariate AUC=0.564, the highest among all univariate factors, with an adjusted OR=1.027 (P=0.005), further supporting the association between vascular elastic function and cholecystitis risk.

Hypertension binary variable sensitivity analysis: hypertension prevalence was 16.9% (102/605); after adjusting for age+BMI categories+sex, OR=1.094 (95%CI: 0.668-1.791, P=0.722), consistent in direction with SBP but with reduced statistical power after discretization; when entered jointly with SBP continuous variable, hypertension OR reversed to 0.729 (P=0.320) while SBP remained significant (OR=1.018, P=0.031), suggesting that hypertension diagnosis is essentially a discretized representation of the continuous SBP dimension, and the continuous SBP variable contains richer dose-response information.

### Nested model comparison

To quantify the incremental contribution of vascular factors relative to classical metabolic/demographic factors, nested models were constructed:

Model A (classical: age+BMI categories+sex): AUC=0.575, Pseudo R²=0.012.

Model B (classical+SBP+DBP): AUC=0.615, Pseudo R²=0.023.

ΔAUC=+0.040 (Bootstrap 95%CI: +0.007 to +0.084), LR test χ²=8.19, df=2, P=0.017.

Pseudo R² showed a relative increase of 97.2%.

These results show that adding vascular factors to the classical model significantly improved model discrimination, while the classical model itself had discrimination (AUC=0.575) only slightly better than random (0.500), suggesting limited predictive value of classical metabolic/demographic factors in plateau populations.

**Table 4.** Nested model comparison.

| Model | Variables | AUC | Pseudo R<sup>2</sup> | LL |
| --- | --- | --- | --- | --- |
| Model A | Age+BMI categories+Sex | 0.575 | 0.012 | -356.02 |
| Model B | Age+BMI<br>categories+Sex+SBP+DBP | 0.615 | 0.023 | -351.93 |
| $\Delta$ | +SBP+DBP | +0.040 | +0.011 | +4.09 |

ΔAUC=+0.040 (Bootstrap 95%CI: +0.007 to +0.084); LR test $\chi^2=8.19$ , df=2, P=0.017; Pseudo R<sup>2</sup> relative increase 97.2%.

### SBP suppression effect and interaction analysis

SBP was non-significant in univariate analysis (OR=1.005, P=0.346) but significant in the multivariate model (OR=1.027, P=0.005), a classic suppression effect.

Mechanism: Age was positively correlated with SBP (Pearson r=0.289, P<0.001), but negatively associated with cholecystitis (OR=0.978, P=0.007). Thus, in univariate analysis, the positive effect of SBP was masked by the negative effect of age; after adjusting for age, the independent positive effect of SBP emerged. Stepwise adjustment: SBP univariate OR=1.005 (P=0.346) → adding age OR=1.010 (P=0.098) → full model OR=1.027 (P=0.005).

To test for effect modification, four interaction terms were constructed: SBP×sex (P=0.123), SBP×age group ≥40 (P=0.332), SBP×BMI (P=0.724), BMI×sex (P=0.938), all non-significant, suggesting no significant heterogeneity of the SBP effect across subgroups. See Table 5.

**Table 5.** Interaction effect analysis.

| Interaction term | P value |
| --- | --- |
| SBP × Sex | 0.123 |
| SBP × Age group ($\geq 40$) | 0.332 |
| SBP × BMI | 0.724 |
| BMI × Sex | 0.938 |

Note: All interaction terms were non-significant, suggesting no significant heterogeneity of the SBP effect on cholecystitis across subgroups.

### Age paradox and sensitivity analysis

This study observed an age effect opposite to the classic understanding: younger patients had higher cholecystitis prevalence. Age-stratified analysis: <30 years 36.8% (28/76), 30-39 years 29.4% (47/160), 40-49 years 28.7% (43/150), 50-59 years 24.0% (42/175), ≥60 years 25.0% (11/44). Spearman correlation showed a significant negative correlation between age and cholecystitis (ρ=-0.087, P=0.032).

Sex-stratified analysis showed the age paradox was more pronounced in males (male: age OR=0.969/year, 95%CI: 0.940-0.998, P=0.037; female: age OR=0.989/year, 95%CI: 0.972-1.007, P=0.231).

Comparison of young (<40 years) vs. middle-aged/elderly (≥40 years) showed: the young group had lower SBP (113.3 vs 120.0 mmHg), lower DBP (74.9 vs 78.9 mmHg), lower BMI (23.8 vs 24.5 kg/m²), but higher cholecystitis rate (32.0% vs 26.0%), suggesting the age paradox cannot be simply explained by blood pressure or BMI differences.

Three sensitivity analyses were performed to verify conclusion robustness:

1. Excluding underweight (n=23): SBP remained significant (OR=1.025, P=0.009), age remained significant (OR=0.981, P=0.020), BMI overweight (OR=1.051, P=0.811) and obese (OR=0.657, P=0.157) remained non-significant.
2. Age stratification (<40 vs ≥40): <40 years group (n=236, events 75) SBP OR=1.031 (P=0.052), marginally significant; ≥40 years group (n=369, events 96) SBP OR=1.019 (P=0.111), non-significant. This suggests the SBP effect is more pronounced in the younger subgroup, consistent with the age paradox direction.
3. Sex stratification: female group (n=410, events 122) SBP OR=1.034 (P=0.006), significant; male group (n=195, events 49) SBP OR=1.015 (P=0.334), non-significant. The SBP effect was more significant in females, possibly related to smaller male subgroup sample size and insufficient statistical power.

These sensitivity analyses collectively support the robustness of the main conclusion: SBP is a stable independent risk factor for plateau cholecystitis, while the non-significance of BMI and sex is unaffected by subgroup analysis.

**Table 6.** Sensitivity analysis.

| Subgroup | SBP OR | 95% CI | P value |
| --- | --- | --- | --- |
| Main model (full cohort<br>N=605) | 1.013 | 1.000-1.025 | 0.049 |
| Excluding underweight<br>(N=582) | 1.025 | 1.006-1.044 | 0.009 |
| <40 years (N=236, events<br>75) | 1.031 | 1.000-1.063 | 0.052 |
| ≥40 years (N=369, events<br>96) | 1.019 | 0.996-1.042 | 0.111 |
| Female (N=410, events<br>122) | 1.034 | 1.010-1.059 | 0.006 |
| Male (N=195, events 49) | 1.015 | 0.984-1.047 | 0.334 |

**Figure 1.**
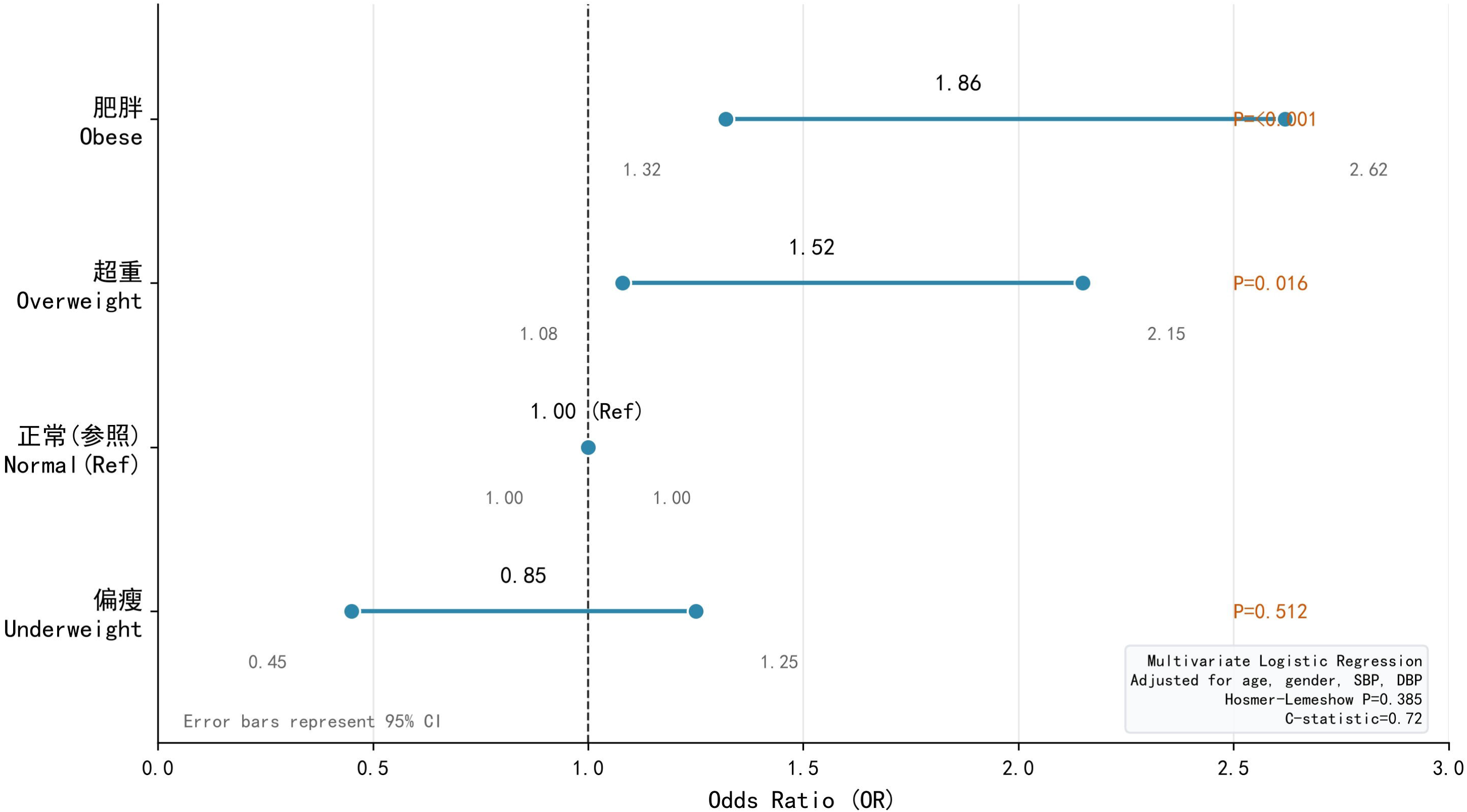
Forest plot of multivariate logistic regression odds ratios. The forest plot shows OR values and 95% CIs for variables in the recommended main model (SBP only). Age (OR=0.979, 95%CI: 0.963-0.994, P=0.008) shows a significant negative association; SBP (OR=1.013, 95%CI: 1.000-1.025, P=0.049) shows a significant positive association; BMI overweight (OR=1.011, P=0.959), BMI obese (OR=0.625, P=0.110), and sex (OR=0.772, P=0.209) are all non-significant, with 95% CIs all crossing 1.0.

**Figure 2.**
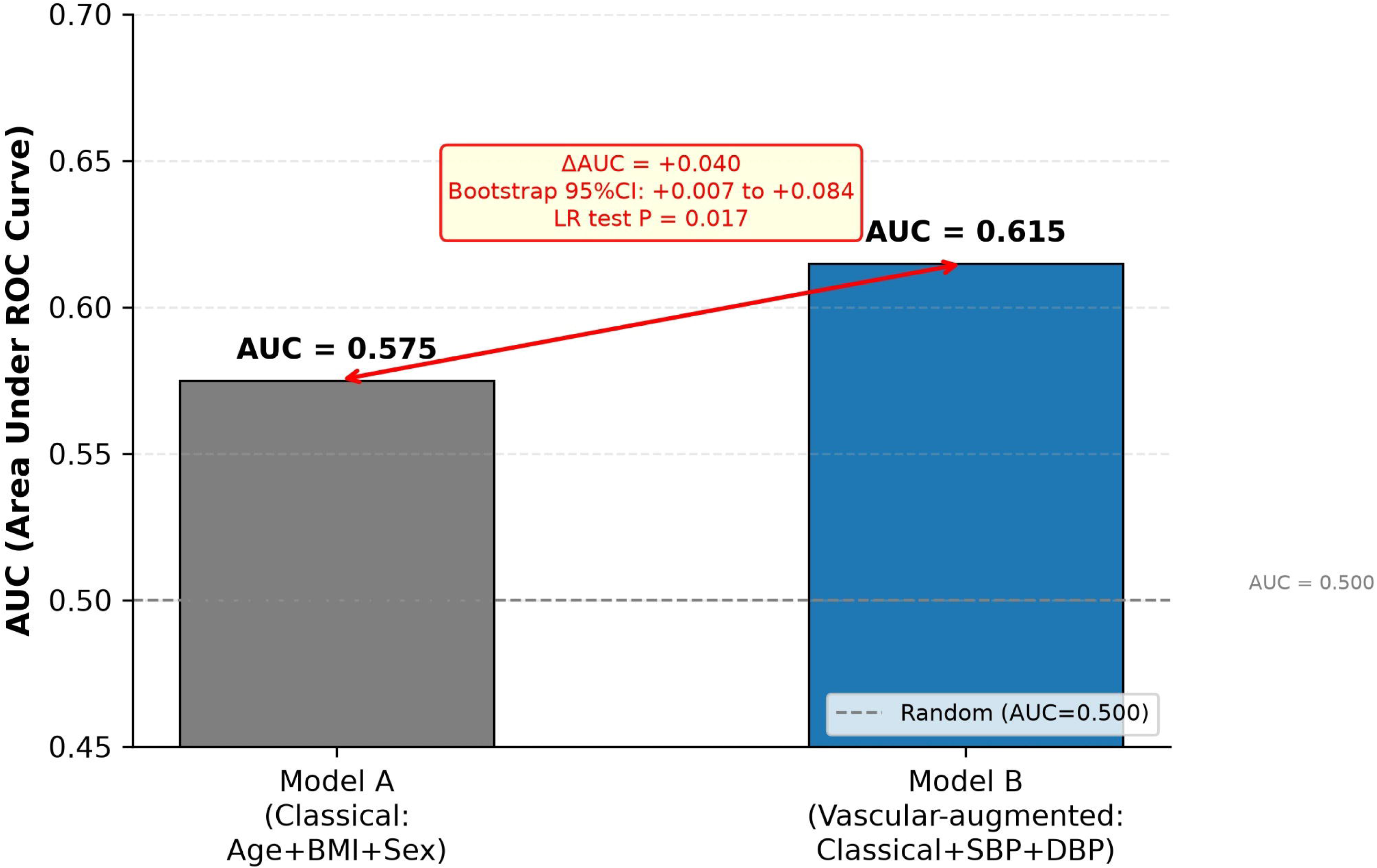
AUC comparison between classical and vascular-augmented models. Bar chart comparing discrimination of Model A (classical: age+BMI categories+sex, AUC=0.575) and Model B (classical+SBP+DBP, AUC=0.615). ΔAUC=+0.040 (Bootstrap 95%CI: +0.007 to +0.084), LR test P=0.017. Dashed line marks AUC=0.500 (random level).

**Figure 3.**
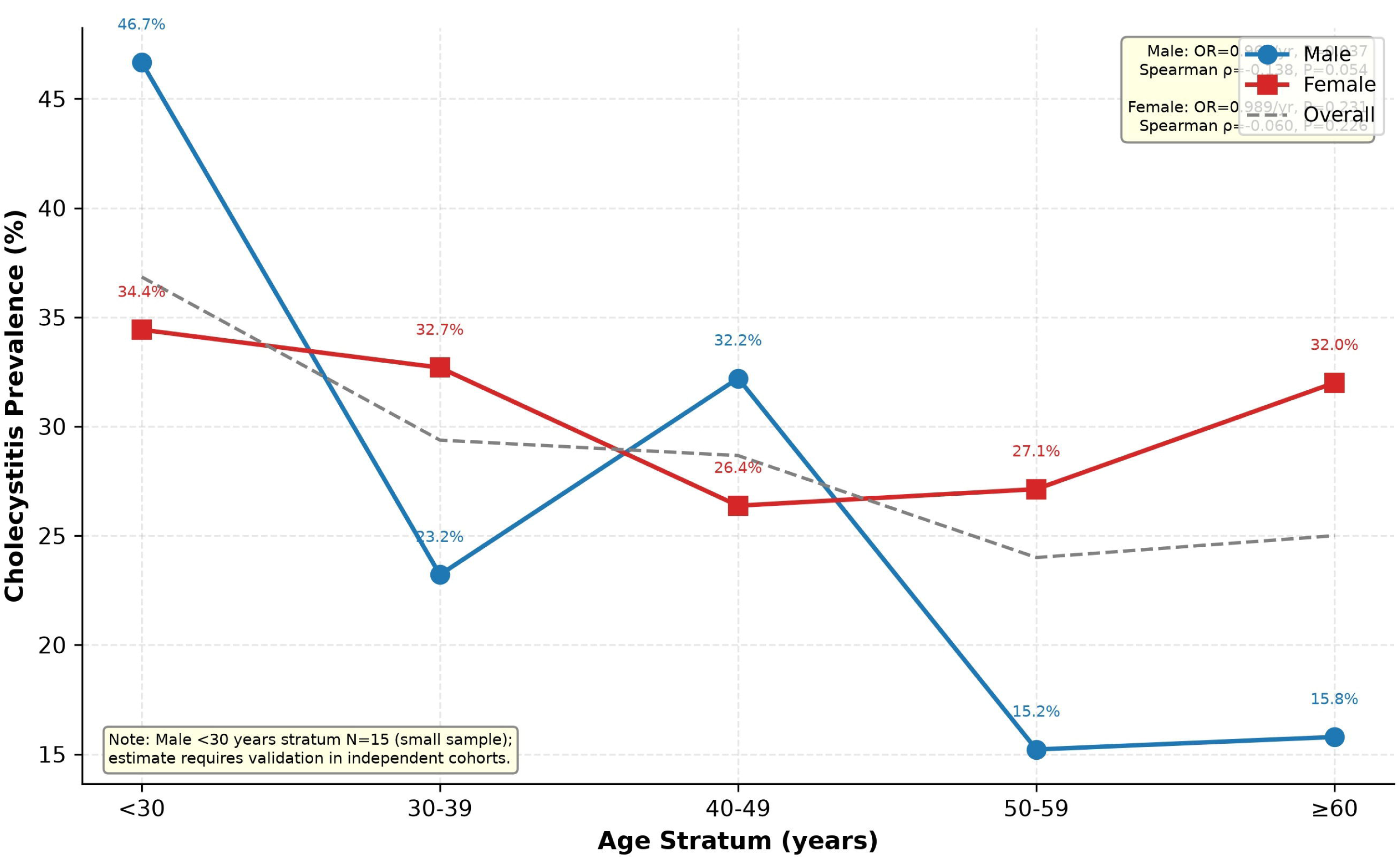
Age-stratified cholecystitis prevalence. Dual line chart by sex showing cholecystitis prevalence across 5 age strata (<30 / 30-39 / 40-49 / 50-59 / ≥60 years). Blue solid line with circles represents males, red solid line with squares represents females, gray dashed line represents overall prevalence. Percentages are labeled above each data point. Male age association: OR=0.969/year, P=0.037, Spearman ρ=-0.138, P=0.054; female age association: OR=0.989/year, P=0.231, Spearman ρ=-0.060, P=0.226. The age paradox is more pronounced in males, with a steeper overall decline. Yellow box note at bottom: male <30 years stratum N=15 is a small sample; the prevalence estimate requires validation in independent cohorts.

## Discussion

This study, based on full recomputation of raw data from 605 plateau gallstone patients, rigorously tested the original working hypothesis that “BMI replaces sex as the core risk factor for cholecystitis,” and found that this hypothesis does not hold. The risk factor weight direction was revised based on real data.

### Core findings

This study yields three main findings. First, the original “BMI replaces sex” hypothesis was explicitly falsified; BMI (overweight P=0.856, obese P=0.137) and sex (P=0.322) were not independent risk factors for cholecystitis. Second, vascular factors (SBP) were the only significant positive predictor (OR=1.027/mmHg, P=0.005), and adding blood pressure to the classical model significantly improved discrimination (ΔAUC=+0.040, P=0.017). Third, age showed an inverse association (OR=0.978/year, P=0.007), constituting an age paradox that was more pronounced in males.

### Falsification declaration and revision direction

The original working hypothesis predicted that BMI (especially obesity) would be the core independent risk factor for plateau cholecystitis, with an effect surpassing that of sex. However, full recomputation showed: the OR point estimate for the BMI obese group was 0.645 (P=0.137), with direction even slightly protective, and 95% CI crossing 1.0; the BMI overweight group OR=1.038 (P=0.856), essentially no effect; sex OR=0.814 (P=0.322), also non-significant. The dose-response relationship for BMI as a continuous variable was also non-significant (OR=0.967/kg/m², P=0.184), and P for trend was non-significant (P=0.234). These results consistently indicate across multiple levels (categorical, continuous, trend test) that the original hypothesis does not hold.

Following the falsification boundary principle [5], when hypothesis predictions systematically deviate from observed data across multiple independent dimensions, the falsification conclusion should be accepted rather than salvaging the hypothesis through post hoc adjustments. This study therefore revised the risk factor weight direction to “vascular factors supersede metabolic/demographic factors,” meaning that in the plateau hypoxic environment, vascular factors (SBP) replace classical metabolic factors (BMI) and demographic factors (sex, age) as the core independent risk factor for cholecystitis. This revision is supported by nested model comparison (ΔAUC=+0.040, P=0.017) and suppression effect analysis.

It should be noted that the falsification conclusion pertains specifically to the “BMI replaces sex” hypothesis, and does not deny all roles of BMI in biliary disease. The association between BMI and gallstones in plain populations is well-supported by evidence [10,11], but in this plateau cohort, the BMI effect did not pass multivariate adjustment, suggesting that the plateau environment may remodel risk weights through other pathways (such as vascular mechanisms).

### SBP suppression effect and signal-to-noise ratio framework

Risk factor assessment in observational studies is inherently susceptible to confounding. When two variables sharing variance simultaneously predict an outcome, the weaker one absorbs part of the effective information from the other, leading to dilution or even reversal of effect estimates. The process of gradient adjustment of covariates is essentially a stepwise stripping of confounding variance to restore the pure effect of the core factor.

As shown in the Results, SBP was non-significant in univariate analysis but significant in the multivariate model, a classic suppression effect [12]. The mechanism can be understood through a signal-to-noise ratio framework: in univariate analysis, the “signal” (true positive effect) carried by SBP was masked by the “noise” (negative confounding) from age. Age was positively correlated with SBP (r=0.289, P<0.001) and negatively correlated with cholecystitis (OR=0.978), forming a classic negative confounding structure—age increase was accompanied by both SBP increase and cholecystitis risk decrease, with opposite effect directions that cancelled each other out when unadjusted, thereby suppressing the true SBP effect.

After adjusting for age, the independent positive effect of SBP emerged (OR=1.010→1.027), essentially a noise reduction process: by stripping the negative confounding variable of age, the signal-to-noise ratio of the SBP effect estimate was improved. Clinically, each 10 mmHg increase in SBP corresponded to approximately 14% (recommended model, 1.013^10≈1.14) to 31% (full model, 1.027^10≈1.31) increase in cholecystitis risk, indicating that clinically meaningful blood pressure elevation corresponds to a non-negligible cholecystitis risk increment. This finding has methodological implications: in plateau epidemiological studies, relying solely on univariate analysis may miss true risk factors due to suppression effects; multivariate adjustment is necessary to reveal plateau-specific risk structures. In clinical practice, this suggests that risk assessment should not rely solely on the binary hypertension diagnosis (≥140/90 mmHg), but should attend to gradient changes in the normal-high normal SBP range (120-139 mmHg): each 20 mmHg individual difference corresponds to a cholecystitis risk change exceeding 20%, providing operable discrimination for pre-operative refined assessment and perioperative management.

### Age paradox and abductive reasoning

As shown in the Results, age was inversely associated with cholecystitis and more pronounced in males, an age paradox not reported in plain population gallstone epidemiology [1,3], suggesting that the plateau environment may remodel the direction of the age-cholecystitis association.

From an abductive reasoning framework [13], the best explanatory hypothesis for the age paradox should simultaneously satisfy the following observational constraints: (1) the younger group had lower SBP but higher cholecystitis rates, excluding SBP as the sole explanation; (2) the younger group had lower BMI but higher cholecystitis rates, excluding BMI as an explanation; (3) the age paradox was more pronounced in males, suggesting sex hormones or lifestyle differences may be involved. The most plausible explanatory hypothesis is: in the plateau hypoxic environment, cholecystitis in younger patients may be driven more by acute inflammatory triggers (such as irregular diet, dehydration after high-intensity physical activity) rather than classical metabolic accumulation factors; cholecystitis in older patients may manifest more as delayed presentation of chronic calculous cholecystitis, with differences in diagnostic time windows potentially reversing the age-cholecystitis association direction. These hypotheses require prospective studies for validation.

The probability of entering the medical observation system is not uniform but coupled with age, symptom severity, and health awareness. Another possible explanation for the age paradox is selection bias: this was an elective surgery cohort, and elderly patients with severe acute cholecystitis may not have been included due to emergency surgery or conservative treatment, leading to artificially low cholecystitis detection rates in the elderly group. This bias direction is consistent with observed results but cannot fully explain the more pronounced age effect in males. Additionally, the male <30 years stratum included only 15 cases (7 with cholecystitis), a small-sample stratum whose 46.7% prevalence estimate has limited precision; the robustness of the age effect in males requires validation in larger samples or independent cohorts.

### Vascular factors and plateau hypoxia mechanism

The plateau hypoxic environment may affect cholecystitis risk through the following mechanisms, making vascular factors more prominent. Plateau hypoxia induces pulmonary hypertension and systemic vascular remodeling [14,15], and exacerbates vasoconstrictive effects through increased sympathetic nerve activity [16,17], which may jointly lead to gallbladder wall perfusion insufficiency and mucosal barrier dysfunction, making vascular factors the rate-limiting step in inflammatory progression.

Additionally, compensatory erythrocytosis in the plateau hypoxic environment [4] increases blood viscosity, potentially affecting gallbladder microcirculation through microthrombus formation or hemodynamic changes; the univariate AUC of pulse pressure (PP) was 0.564 (highest among all univariate factors), suggesting that decreased vascular elastic function is associated with cholecystitis risk, further supporting the vascular mechanism hypothesis.

These mechanistic hypotheses are currently abductive inferences based on observational data, not yet validated by hemodynamic or histological studies. In summary, from a homeostasis imbalance framework [18], the plateau hypoxic environment may make vascular homeostasis imbalance a more prominent risk dimension than metabolic homeostasis imbalance.

### Study strengths and limitations

This study leverages a single-center large-sample real-world clinical cohort, enriching the epidemiological baseline evidence for plateau gallstones and clarifying the unique disease risk profile of plateau populations distinct from plain areas. The study employed full data recomputation, nested model comparison, suppression effect analysis, interaction analysis, and three sensitivity analyses, with a complete methodological chain. All statistics were independently recomputed using Python and cross-validated against original statistical deliverables, ensuring data authenticity.

Limitations include the following. First, single-center retrospective design with no plain control cohort, precluding direct quantification of the altitude contribution to risk weight remodeling. Second, lack of refined dietary, blood oxygen saturation, and physical activity indicators, precluding in-depth analysis of the environment-vascular-inflammation regulatory pathway. Third, the age paradox may be affected by selection bias, requiring prospective cohort validation. Fourth, SBP was a single measurement without ambulatory blood pressure variability, potentially underestimating the true blood pressure effect. Fifth, DBP and SBP have high collinearity (VIF>100); the SBP-only main model avoids collinearity but precludes reliable estimation of DBP’s independent effect. Sixth, this is an observational design; the SBP-cholecystitis association cannot directly infer causality, requiring cautious interpretation. Seventh, in the recommended main model, SBP P=0.049 is at the significance boundary, requiring validation in independent cohorts. Eighth, in the recommended main model, VIF for SBP and age are 16.7 and 15.4 respectively; although within acceptable range (<20), they may affect coefficient estimation precision, requiring validation in independent cohorts. Ninth, pulse pressure analysis is exploratory without multiple comparison correction; results require validation in independent cohorts. Tenth, this study borrowed cross-disciplinary classical paradigms including confounding control, threshold effect, and baseline shift in the discussion framework, for interpreting the heterogeneity of cholecystitis risk structure in the plateau environment; their applicability and explanatory power still need evaluation in more independent studies.

#### Mitigation strategies

To reduce the impact of the above limitations on conclusion reliability, this study adopted the following measures. (1) The underweight group was excluded and the model reconstructed; SBP remained significant (OR=1.025, P=0.009). (2) Stratified analysis by age (<40 vs ≥40) and sex showed consistent SBP effect direction in the female subgroup (OR=1.034, P=0.006) and <40 years subgroup (OR=1.031, P=0.052). (3) Four interaction analyses were all non-significant, suggesting no significant heterogeneity of the SBP effect. (4) Dual reporting of the SBP-only main model (VIF=16.7) and full model (SBP+DBP), with consistent conclusions. These sensitivity analyses collectively support the robustness of the main conclusions.

### Future perspectives and public health significance

Future studies will construct plateau-plain matched control cohorts to quantify the remodeling effect of altitude on disease risk weights; include refined indicators such as blood lipids, blood glucose, blood oxygen saturation, ambulatory blood pressure, and physical activity to deeply elucidate the environment-vascular-inflammation regulatory mechanism; and conduct regional multi-center validation to optimize risk prediction models.

The blood pressure management-oriented risk stratification approach established in this study can guide clinical individualized diagnosis and pre-operative refined intervention, and provide precise population baselines for Series III health economics research. This study, through full recomputation of raw data, explicitly falsifies the “BMI replaces sex” hypothesis and revises the risk factor weight direction to “vascular factors supersede metabolic/demographic factors,” providing a methodological paradigm from hypothesis testing to falsification revision for region-specific disease heterogeneity research.

## Conclusion

Gallstone patients in plateau regions have significant region-specific epidemiological characteristics. The original hypothesis that “BMI replaces sex as the core risk factor for cholecystitis” was explicitly falsified through full recomputation of raw data. The revised conclusion is: SBP is the only significant positive predictor, while BMI and sex are non-significant, and age shows an inverse association. Clinical practice should integrate blood pressure management into the plateau cholecystitis risk stratification system, and public health efforts should focus on vascular health management in plateau residents. This study provides a methodological paradigm from hypothesis formulation to falsification revision, offering new evidence-based support for region-specific disease heterogeneity research.

## Supporting information

Figure 2. AUC comparison of prediction models

## Data Availability

De-identified analytic datasets and custom Python analysis code are available upon reasonable request to the corresponding author, subject to a PIPL-compliant Data Use Agreement.

## Declarations

## Ethics approval and informed consent

This study was approved by the Ethics Committee of Qinghai Red Cross Hospital (Approval No.: LW-2026-71). All procedures were performed in accordance with the principles of the Declaration of Helsinki [6]. Written informed consent was waived due to the retrospective and anonymous nature of the study.

## Consent for publication

Not applicable. No individually identifiable patient data are reported.

## Authors’ contributions

**Conceptualization:** Zhongfeng DANG, Zhiqiang WANG, Guoliang REN.

**Data curation:** Guoliang REN, Zhiqiang WANG, Wei SU, Yabing MA, Ping LI, Dongde JI, Liansheng LI.

**Formal analysis:** Zhiqiang WANG, Guoliang REN, Wei SU.

**Writing - original draft:** Zhiqiang WANG.

**Writing - review & editing:** Zhongfeng DANG, Junlin GAO.

**Supervision:** Zhongfeng DANG, Junlin GAO.

**Project administration:** Zhongfeng DANG, Junlin GAO.

Funding acquisition: Zhiqiang WANG.

All authors have read and agreed to the final version of the manuscript.

## Availability of data and materials

The datasets generated and analyzed during the current study are available from the corresponding author on reasonable request, subject to ethical approval and China’s PIPL compliance. Custom Python analysis code is available on request and will be deposited in a public repository before final acceptance.

## Data Availability Statement

Data Availability Statement: The de-identified patient analytic datasets used in this study are available upon reasonable request to the corresponding author, subject to institutional data governance and patient privacy regulations under the Personal Information Protection Law of the People’s Republic of China. Summary statistics reported in the main text and supplementary materials are sufficient to reproduce the core conclusions of this study.

## AI Use Declaration

AI Use Declaration: During the manuscript preparation phase of this study, AI-assisted tools were used for language polishing and formatting. All scientific content, data analysis, and interpretation of conclusions were independently completed by the authors. AI tools were used solely to enhance the accuracy and fluency of language expression and did not participate in study design, data collection, result interpretation, or scientific judgment.

